# Accuracy of the National Early Warning Score version 2 (NEWS2) in predicting need for time-critical treatment: Retrospective observational cohort study

**DOI:** 10.1101/2024.09.11.24313201

**Authors:** Steve Goodacre, Laura Sutton, Gordon Fuller, Ashleigh Trimble, Richard Pilbery

## Abstract

**Background:** Initial emergency department (ED) assessment can use early warning scores to identify and prioritise patients who need time-critical treatment. We aimed to determine the accuracy of the National Early Warning Score version 2 (NEWS2) for predicting the need for time-critical treatment.

**Methods:** We undertook a single centre retrospective observational cohort study. We randomly selected 4000 adults who attended a tertiary hospital ED in 2022 and had NEWS2 routinely recorded on electronic patient records. The first NEWS2 score and vital signs were extracted from electronic records. Research nurses selected cases that received a potentially time-critical treatment. Two independent clinical experts then determined whether time-critical treatment was or should have been received, using an expert consensus derived list of interventions. We used receiver operating characteristic (ROC) analysis and calculated sensitivity and specified at pre-defined thresholds to evaluate the accuracy of NEWS2 for predicting need for time-critical intervention.

**Results:** After excluding ten patients who received their intervention before NEWS2 recording, 164/3990 (4.1%) needed time-critical treatment and 71/3990 (1.8%) died within seven days. NEWS2 predicted need for time-critical treatment with a c-statistic of 0.807 (95% confidence interval 0.765 to 0.849) and death within seven days with a c-statistic of 0.865 (0.813, 0.917). NEWS2>4 predicted need for time-critical treatment with sensitivity of 0.518 (0.442, 0.593) and positive predictive value of 0.258 (0.213, 0.307). Patients needing emergency surgery, antibiotics for open fractures, insulin infusion, or manipulation of limb-threatening injuries frequently had NEWS2≤4. Patients with NEWS2>4 who did not need time-critical treatment frequently scored three points on NEWS2 for their respiratory rate, conscious level, or receiving supplemental oxygen.

**Conclusion:** NEWS2 has modest accuracy for predicting need for time-critical treatment. We have identified time-critical interventions that frequently have low NEWS2 scores and NEWS2 parameters than may overestimate need for time-critical intervention.

## Background

Patients attending the emergency department (ED) can face frequent and sometimes prolonged waits before they receive definitive assessment and treatment. This can result in avoidable harm if urgent treatment is delayed. Initial assessment of patients arriving at the ED aims to reduce this risk by ensuring that patients with time-critical conditions are identified and prioritised [1,2].

Early warning scores use physiological measurements to produce a composite score reflecting illness severity that can assist initial assessment. In the United Kingdom (UK), guidance from the Royal College of Emergency Medicine supports the use of early warning scores as part of initial assessment but advises against their use as a sole measure of acuity [1]. The Royal College of Physicians (RCP) developed the National Early Warning Score version 2 (NEWS2) to standardise the assessment and response to acute illness in adults and recommends that ED staff use NEWS2 to aid the initial assessment of adult patients [3]. NHS England has endorsed the use of NEWS2 and provided guidance to support adoption in acute and ambulance settings [4].

Systematic reviews and meta-analysis have shown that early warning scores have good prediction for mortality but adequate to poor prediction of intensive care or hospital admission [5-8]. Mortality reflects illness severity, including frailty and long-term conditions, but early warning scores need to predict illness acuity – the need for time-critical treatment. Mortality identifies deaths that occurred despite treatment, and may therefore have been inevitable, but does not identify cases where treatment prevented death [9]. Mortality may therefore fail to identify cases most likely to benefit from time-critical treatment. A recent review of ED acuity assessment tools [10] identified only one small study that directly measured accuracy for time-critical treatment [11] and a systematic review that included studies measuring accuracy for time-critical diagnoses [12].

We aimed to determine the accuracy with which NEWS2 predicts the need for time-critical treatment among adults attending the ED and characterise presentations where NEWS2 has poor accuracy.

## Methods

We undertook a single centre retrospective observational cohort study at the Northern General Hospital ED in Sheffield, UK. This is the only adult ED serving the 530,000 population of Sheffield and the adult major trauma centre for the 1.8 million population of South Yorkshire. The ED receives all undifferentiated adult emergencies except ambulance arrivals with stroke or ST-elevation myocardial infarction requiring reperfusion, which are taken directly to specialist services. At initial assessment nurses record vital signs on the ED information system for all patients considered to be at risk of physiological deterioration, which then generates a NEWS2 score for patients with a complete set of vital signs. Oxygen saturation is recorded using scale 1 unless the patient is definitely known to have hypercapnic respiratory failure. Patients with minor injuries or primary care complaints may be referred to services located alongside the ED. The attending clinician completes a coding form when ED assessment is complete that records standardised ED diagnoses and treatments.

We used routine hospital data to identify all adult (aged 16 or over) ED attendances in 2022 that had a NEWS2 score recorded and randomly sampled 4000 attendances to account for seasonality, having excluded repeat attendances and patients who had opted out of allowing their data to be used for research. We extracted the following routine hospital data: age, sex, ethnicity, the first recorded NEWS2 score, heart rate, respiratory rate, temperature, peripheral oxygen saturation, blood pressure and conscious level, ED diagnoses and treatments, hospital admission, and attendances, admissions, and deaths over the following week.

A research nurse reviewed the ED records, initial receiving records for admitted patients, and hospital discharge summaries of the selected attendances to identify patients who received a potentially time-critical treatment or suffered an adverse outcome (death or safety incident) that could have been prevented by time-critical treatment. Two independent experts then reviewed hospital records of the selected patients to determine whether they had received or should have received a time-critical treatment using the definition outlined below, with a third expert resolving any disagreements. The experts (SG, AT, GF) had all completed specialist training in emergency medicine. The NEWS2 score was recorded on an observations chart that was not part of the hospital records reviewed by the experts, so outcome adjudication was undertaken by observers who were not aware of the NEWS2 score but could have estimated or calculated the score from the clinical observations.

We also checked all the selected attendances against the ED database of patient safety incidents recorded using the DATIX system to identify any adverse events that could have been prevented by time-critical treatment. The incident reports were used to select any potentially relevant incidents, which were then independently adjudicated by two of the clinical experts.

The primary outcome, need for time-critical treatment, was defined through the expert consensus process described in supplementary appendix 1. Nine experts in emergency medicine used existing literature and their clinical experience to define 34 interventions as likely to be time-critical and provided advice on how the list of interventions should be used in outcome adjudication. The clinical experts then used this list of interventions to determine whether each patient had appropriately received or should have received a time-critical treatment.

We excluded cases from the analysis if they received a time-critical treatment in the ED before NEWS2 was recorded or received continuing time-critical prehospital treatment in the ED (for example, airway or breathing support that was initiated prehospital and continued in the ED). We included cases that had received time-critical prehospital intervention if the intervention had been completed and NEWS2 was recorded after completion of the prehospital intervention (for example, treated hypoglycaemia).

A medical statistician from the University of Sheffield (LS) undertook all analysis. NEWS2 scores generated by the hospital information system were verified against their constituent elements. The information system only generates a NEWS2 score if all constituent elements are entered and sets limits for the values entered, so there were no missing data in the study population. However, the system allocates a NEWS2 score of three for any variable with a zero value. We therefore checked any zero values against the hospital records to ensure that the score of three was appropriately allocated (e.g. unrecordable temperature due to hypothermia or unrecordable blood pressure due to shock) and imputed the next available measurement if the zero value appeared to be due to equipment failure.

We undertook receiver operating characteristic (ROC) analysis to determine the discriminant value of NEWS2 for predicting the need for urgent treatment across varying thresholds [13]. We repeated this analysis using a secondary outcome of death within seven days and a composite secondary outcome of death within seven days or need for time-critical treatment, and tested the hypothesis that NEWS2 prediction differs between time-critical treatment and death. We calculated the sensitivity, specificity, positive predictive and negative predictive values (with 95% confidence intervals calculated using the Wilson score method [14]) for thresholds of NEWS2>4 and NEWS2>6. We repeated this analysis with the score classified as being above the threshold if any NEWS2 parameter equals three, in accordance with RCP guidance [3]. Finally, we described the characteristics of the ‘false negative’ cases that had NEWS2≤4 and needed time-critical treatment, and the ‘false positive’ cases that had NEWS2>4 but did not need time-critical treatment.

We estimated that a sample size of 4000 would include 200 cases with the primary outcome. This sample would give an estimated 95% confidence interval (CI) of 0.73 to 0.81 for an assumed c-statistic of 0.77, [13] based on a previous similar study [15].

### Patient and public involvement

Sheffield Emergency Care Forum is a patient and public representative group with an interest in emergency care that has extensive experience of involvement in emergency care research [16]. Two members of the forum joined the research team and were specifically responsible for reviewing the list of time-critical interventions to ensure that it reflected public values and would not discriminate against any patient group.

### Ethical approval

The Health Research Authority and Health and Care Research Wales approved the study (reference 23/HRA/4572).

## Results

Supplementary figure 1 shows the flow of cases through the study. There were 85499 first and 41220 repeat ED attendances in 2022, with 56145/85499 (65.7%) first attendances having NEWS2 recorded. The 27905 patients without NEWS2 at first attendance were relatively young (mean age 44.5 years), with a low admission rate (1911/27905, 6.8%) and relatively high proportions of minor injuries (14346/27905, 46%) and referral to primary care (5423/27905, 17.4%). We excluded 2689 eligible cases that had opted out of allowing data use and 46 aged <16 years, and then randomly selected 4000 from 53410 eligible attendances for inclusion.

The research nurses selected 704/4000 cases with possible time-critical interventions for expert review, with 173/704 adjudicated as needing time-critical treatment (κ=0.89, 95% CI 0.86, 0.93). One additional case was identified through safety incident review, and ten cases were excluded because the time-critical intervention was received before NEWS2 was recorded. Therefore 164/3990 (4.1%) cases were positive for the primary outcome. There were 71 participants (1.8%) experiencing the secondary outcome of death within seven days and 195 (4.9%) with the composite secondary outcome. Table 1 shows the characteristics of the included patients and table 2 shows the time-critical interventions received. The most frequent intervention was IV antibiotics for infection causing new organ dysfunction or shock (66/164).

**Table 1:**
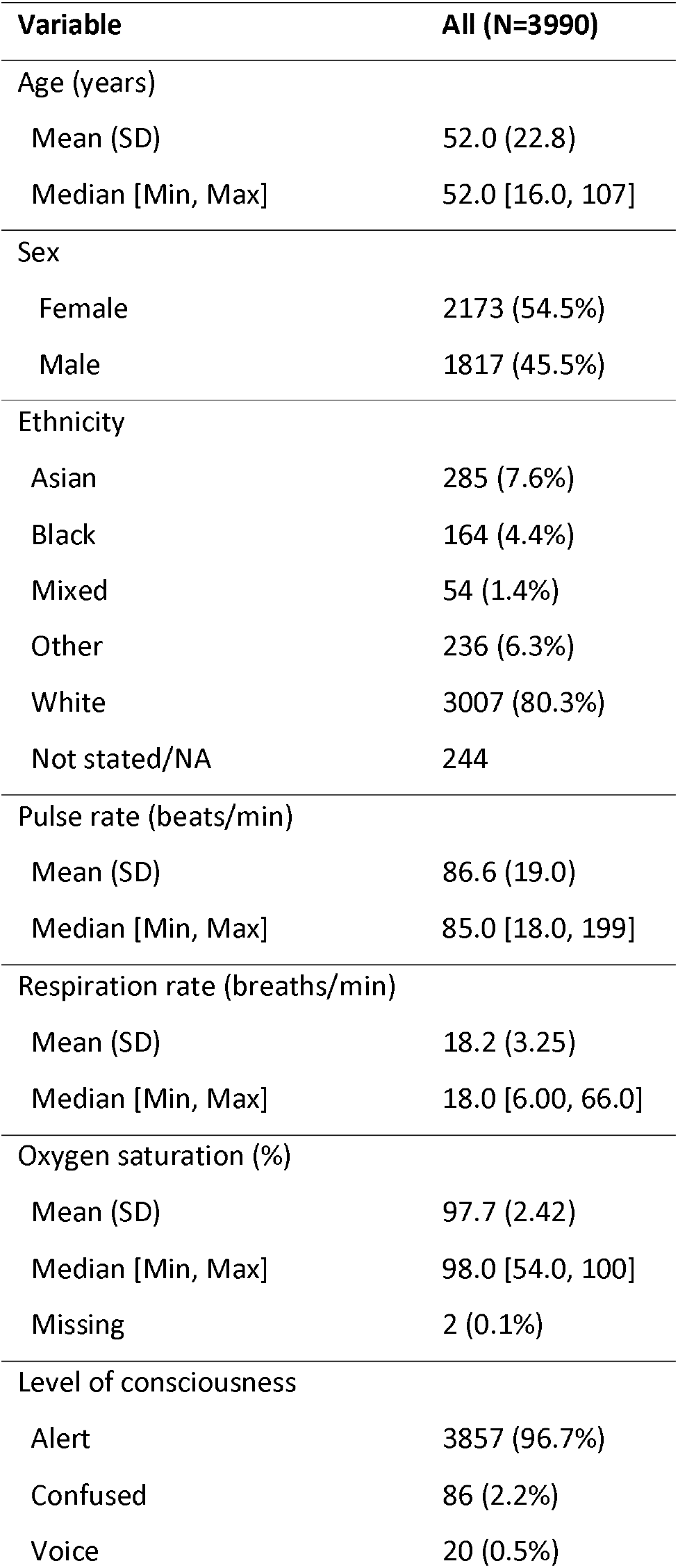
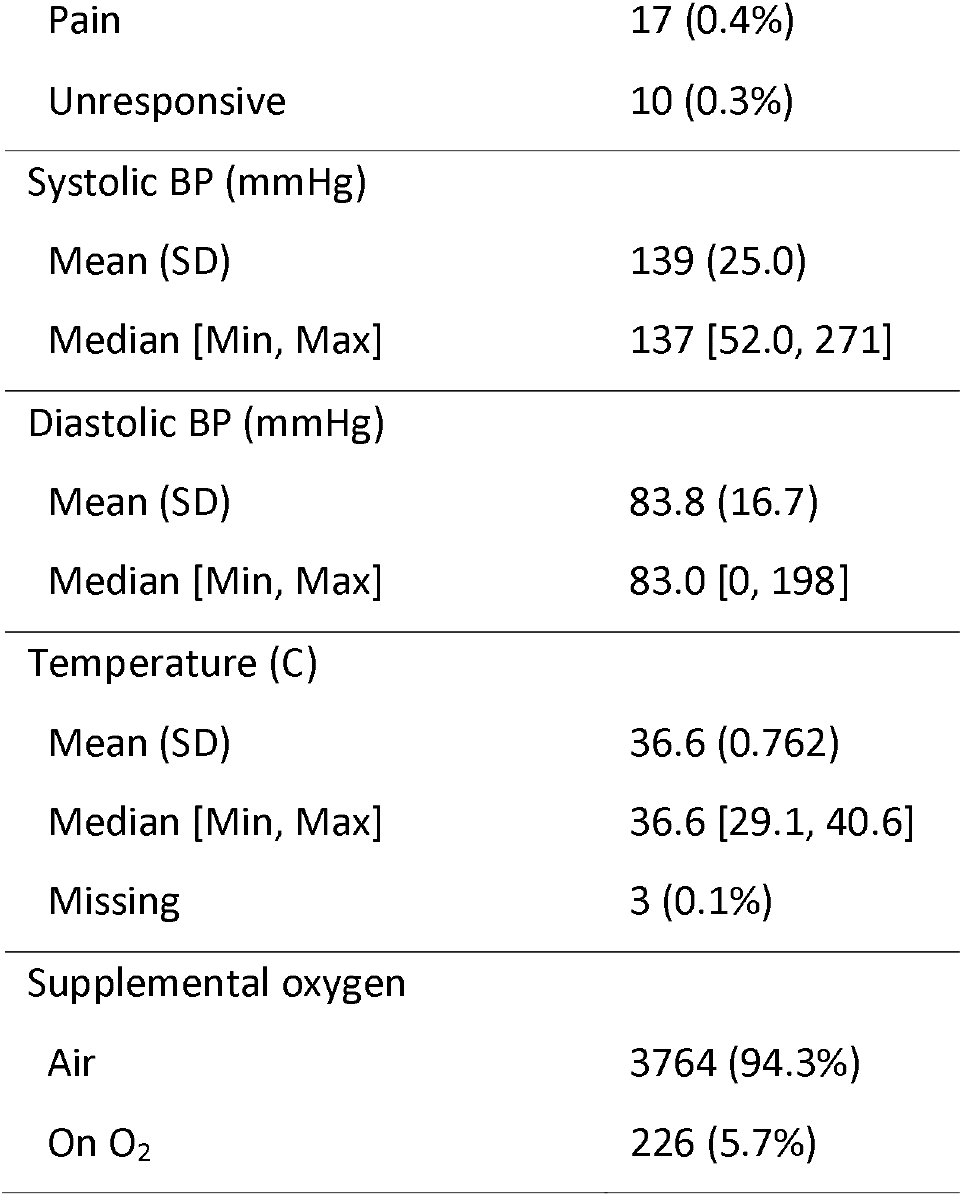
Participant characteristics (whole cohort)

**Table 2:** Treatment received or should have been received in reference standard positive cases.

| **Intervention** | **Frequency** |
| --- | --- |
| IV antibiotics for infection causing new organ dysfunction or shock | 66 |
| Emergency surgery to avoid death or significant disability, including surgical source control for infection | 21 |
| Large volume IV fluid replacement (>2L within 2 hours or >1L with central venous monitoring) | 19 |
| Any intervention to support ventilation (except supplemental oxygen), other during sedation for a procedure | 18 |
| Administration of blood products for acute blood loss, to allow emergency procedures, in haematological emergencies, or severe anaemia in context of proven acute coronary syndrome | 15 |
| Unplanned airway intervention to provide a patent airway, other than during sedation for a procedure | 11 |
| IV antibiotics for genuine open fractures | 11 |
| Hyperkalaemia or hypokalaemia involving IV treatment and cardiac monitoring | 10 |
| Insulin infusion as part of treatment protocol for diabetic ketoacidosis or hyperosmolar hyperglycaemic state | 8 |
| Antidote for life or disability threatening poisoning | 7 |
| Any intervention to support circulation (including CPR), other than intravenous fluids | 7 |
| IV fluids and steroids for Addisonian crisis | 6 |
| Reduction of limb-threatening fracture or dislocation (including threat to skin, nerve or perfusion) | 5 |
| Cardioversion or rate control for life-threatening arrhythmia | 3 |
| Primary percutaneous coronary intervention or thrombolysis for myocardial infarction | 3 |
| Parenteral treatment for hypoglycaemia | 3 |
| Hypertonic saline for hyponatraemia causing a specific neurological disturbance, such as seizures or reduced conscious level | 2 |
| Any intervention to achieve haemorrhage control, other than manual pressure or a dressing | 2 |
| IV nitrates for acute heart failure | 2 |
| IV antibiotics for meningitis or necrotising fasciitis | 2 |
| Emergency reperfusion of an ischaemic limb or organ (e.g. testicular torsion) | 2 |
| Active rewarming for hypothermia | 1 |
| IV treatment to lower life or disability threatening blood pressure | 1 |
| Thrombolysis for massive pulmonary embolism | 1 |
| Any other, not previously specified – acyclovir for herpes simplex meningitis | 1 |

Figure 1 shows the proportion of cases needing time-critical treatment at each NEWS2 score. The proportion increases with NEWS2 score and exceeds a quarter of cases at NEWS2=7 and half of cases at NEWS2=10, albeit based on small numbers at higher NEWS2 score. Figure 2 shows the ROC curves for the primary and secondary outcomes. The c-statistic for NEWS2 prediction the need for time-critical intervention was 0.807 (95% CI 0.765 to 0.869), which was lower than the c-statistic for death within seven days (0.865, 95% CI 0.813, 0.917), although the difference was not statistically significant (p=0.09).

**Figure 1.**
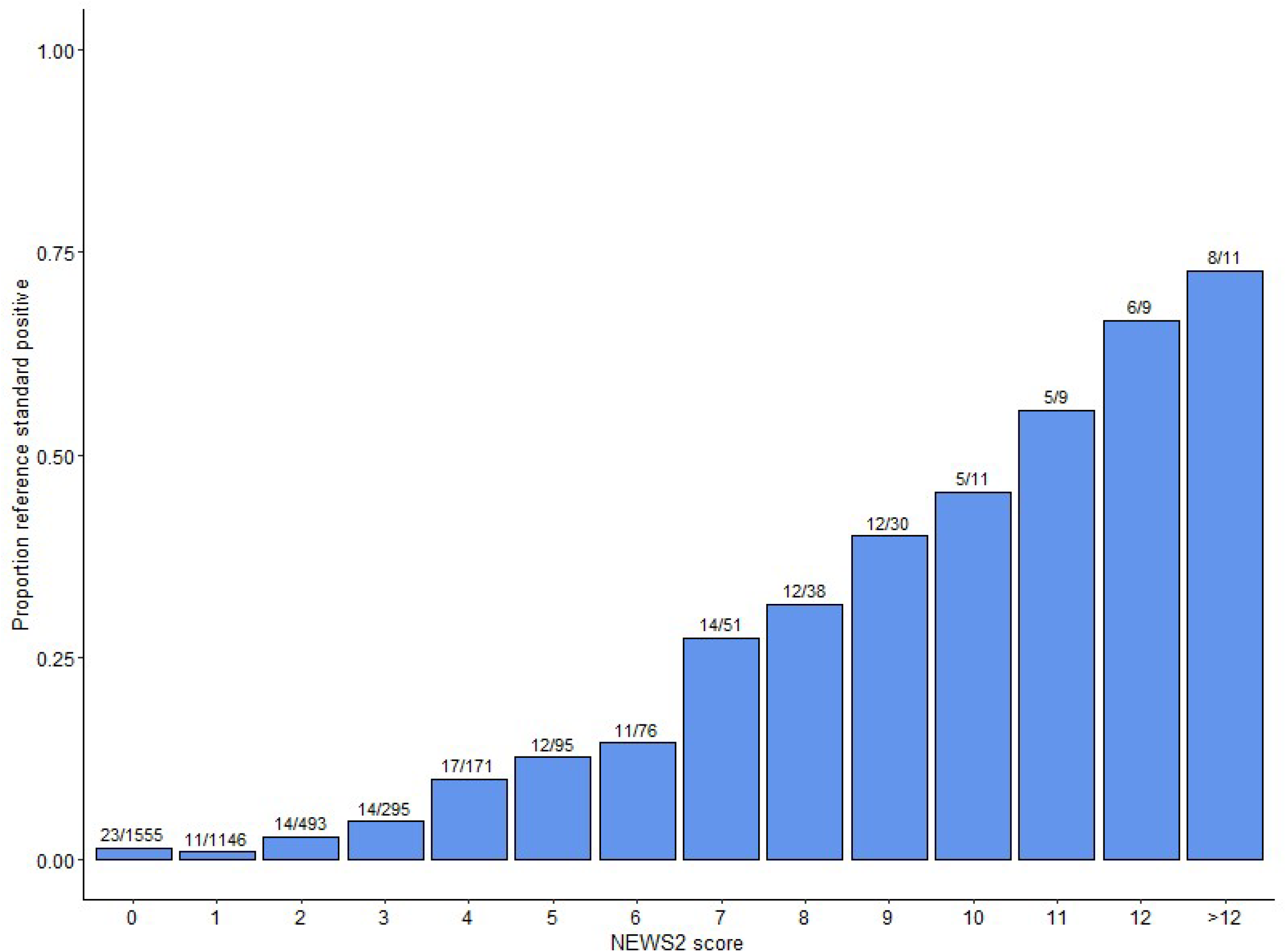
Proportion of cases needing time-critical treatment at each NEWS2 score. The numbers comprising the proportion are shown above each column.

**Figure 2.**
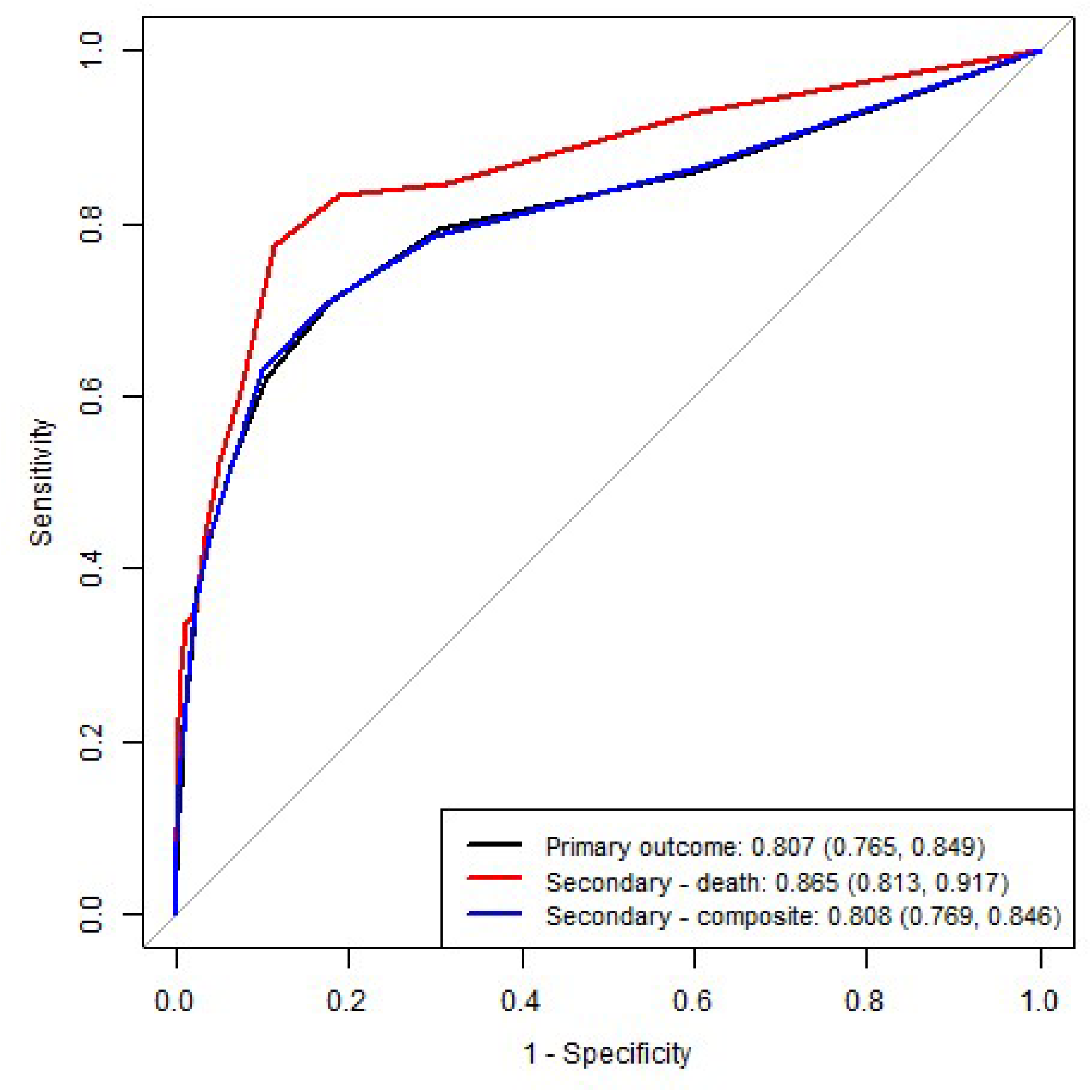
AUROC (95% CI) for NEWS2 and primary and secondary outcomes.

Table 3 shows the accuracy of NEWS2 for the primary outcome using recommended thresholds for patient prioritisation. Around half of patients needing time-critical intervention have NEWS2>4 and around a quarter with NEWS2>4 need time-critical intervention. Sensitivity can be improved to around 60%, at the expense of specificity, by including those with any NEWS2 parameter equalling three as index test positive.

**Table 3:** Diagnostic accuracy statistics (95% CI) for the primary outcome.

| EWS threshold | TP | FP | FN | TN | Sensitivity | Specificity | PPV | NPV | LR+ | LR- |
| --- | --- | --- | --- | --- | --- | --- | --- | --- | --- | --- |
| NEWS2 > 4 | 85 | 245 | 79 | 3581 | 0.518<br>(0.442, 0.593) | 0.936<br>(0.928, 0.943) | 0.258<br>(0.213, 0.307) | 0.978<br>(0.973, 0.983) | 8.094<br>(6.687, 9.796) | 0.515<br>(0.439, 0.603) |
| NEWS2 > 4 or<br>parameter 3 | 98 | 428 | 66 | 3398 | 0.598<br>(0.521, 0.67) | 0.888<br>(0.878, 0.898) | 0.186<br>(0.155, 0.222) | 0.981<br>(0.976, 0.985) | 5.342<br>(4.579, 6.232) | 0.453<br>(0.376, 0.546) |
| NEWS2 > 6 | 62 | 97 | 102 | 3729 | 0.378<br>(0.307, 0.454) | 0.975<br>(0.969, 0.979) | 0.39<br>(0.318, 0.467) | 0.973<br>(0.968, 0.978) | 14.911<br>(11.295, 19.685) | 0.638<br>(0.566, 0.719) |
| NEWS2 > 6 or<br>parameter 3 | 88 | 380 | 76 | 3446 | 0.537<br>(0.46, 0.611) | 0.901<br>(0.891, 0.91) | 0.188<br>(0.155, 0.226) | 0.978<br>(0.973, 0.983) | 5.403<br>(4.552, 6.412) | 0.515<br>(0.436, 0.607) |

Table 4 shows the time-critical interventions that the false negative cases with NEWS2≤4 needed. Patients needing emergency surgery (18/21), IV antibiotics for open fractures (9/11), insulin infusion (5/8), and reduction of limb-threatening injury (5/5) often had NEWS2≤4. The frequency of IV antibiotics for infection causing new organ dysfunction or shock among false negative cases reflects the high frequency of this time-critical intervention (14/66).

**Table 4:** Time-critical treatments that patients with NEWS2≤4 needed (N=79)

| **Intervention** | **Frequency** |
| --- | --- |
| Emergency surgery to avoid death or significant disability, including surgical source control for infection | 18 |
| IV antibiotics for infection causing new organ dysfunction or shock | 14 |
| IV antibiotics for genuine open fractures | 9 |
| Administration of blood products for acute blood loss, to allow emergency procedures, in haematological emergencies, or severe anaemia in context of proven acute coronary syndrome | 7 |
| Insulin infusion as part of treatment protocol for diabetic ketoacidosis or hyperosmolar hyperglycaemic state | 5 |
| Large volume IV fluid replacement (>2L within 2 hours or >1L with central venous monitoring) | 5 |
| Reduction of limb-threatening fracture or dislocation (including threat to skin, nerve or perfusion) | 5 |
| Antidote for life or disability threatening poisoning | 5 |
| Hyperkalaemia or hypokalaemia involving IV treatment and cardiac monitoring | 4 |
| IV fluids and steroids for Addisonian crisis | 4 |
| Any intervention to support ventilation (except supplemental oxygen), other during sedation for a procedure | 4 |
| Cardioversion or rate control for life-threatening arrhythmia | 3 |
| Primary percutaneous coronary intervention or thrombolysis for myocardial infarction | 3 |
| Unplanned airway intervention to provide a patent airway, other than during sedation for a procedure | 2 |
| Any intervention to achieve haemorrhage control, other than manual pressure or a dressing | 2 |

| Intervention | Frequency |
| --- | --- |
| Any intervention to support circulation (including CPR), other than intravenous fluids | 2 |
| IV nitrates for acute heart failure | 2 |
| Emergency reperfusion of an ischaemic limb or organ (e.g. testicular torsion) | 2 |
| Parenteral treatment for hypoglycaemia | 2 |
| Hypertonic saline for hyponatraemia causing a specific neurological disturbance, such as seizures or reduced conscious level | 1 |
| IV treatment to lower life or disability threatening blood pressure | 1 |
| IV antibiotics for meningitis or necrotising fasciitis | 1 |
| Thrombolysis for massive pulmonary embolism | 1 |
| Any other, not previously specified | 1 |

Supplementary table 1 shows the characteristics of the false negative cases and supplementary table 2 shows the characteristics of the false positive cases with NEWS2>4 who did not need time-critical treatment. Supplementary table 3 shows the frequency of NEWS2 parameters scoring three in the false positive cases. Respiratory rate, supplemental oxygen, and altered consciousness frequently contributed three points in false positive cases.

## Discussion

We found that NEWS2 had modest accuracy in predicting need for time-critical treatment among adult ED attenders. The c-statistic for predicting the need for time-critical treatment was 0.807 (95% CI 0.765 to 0.849). Using a threshold of NEWS2>4 would fail to predict around half of cases needing time-critical intervention and three-quarters of patients with NEWS2>4 would not need time-critical intervention. Most patients needing emergency surgery, antibiotics for open fractures, insulin infusion, or manipulation of limb-threatening injuries had NEWS2≤4. Patients with NEWS2>4 who did not need time-critical treatment frequently scored three points for respiratory rate, supplemental oxygen, or conscious level on NEWS2.

Previous studies have shown that early warning scores have better accuracy for predicting death than for predicting hospital or intensive care admission [5-8]. Meta-analysis of nine studies of NEWS in ED patients reported c-statistics of 0.88, 0.86 and 0.77 for 24-hour, 48-hour, and in-hospital mortality, 0.68 for hospital admission, and 0.69 for intensive care admission [5]. A systematic review of 22 studies in acute medical units reported c-statistics of 0.7 to 0.9 for mortality and <0.6 for intensive care admission [6]. Our findings show similar prediction of mortality to previous studies.

Few studies have examined accuracy for time-critical treatment. Hong et al [11] compared the Emergency Severity Index and Simple Triage and Rapid Treatment triage tool in predicting 21/233 cases requiring emergent intervention. Hinson et al [12] systematically reviewed triage systems and identified a few studies that evaluated prediction for specific time-critical diagnoses, such as sepsis and myocardial infarction, but none that evaluated accuracy for time-critical interventions.

Our study has shown that it is possible to measure the accuracy of early warning scores for predicting need for time-critical treatment. We developed a reproducible method for adjudicating the primary outcome based on expert consensus and previous literature that we implemented with excellent inter-observer agreement. Our study also had low rates of missing data due to the ability of the research team to access hospital records and a sample size that allowed accuracy to be estimated with reasonable precision.

Our study had limitations that need to be appreciated. The outcome adjudicators were not aware of the NEWS2 score but knowledge of the observations that comprise the NEWS2 could have influenced their judgements. We only used the first recorded NEWS2 score, whereas repeated scores may provide more information, although this may not be compatible with brief initial assessment. The analyses of false negative and false positive cases were based on small numbers of cases. Our definition of time-critical intervention may be contested, with a large proportion of cases involving IV antibiotics for infection causing new organ dysfunction or shock, which is based on limited evidence [17]. Greater use of scale 2 to record oxygen saturation for patients with potential rather than definite hypercapnic respiratory failure could reduce the number of false positives arising from low oxygen saturation. Finally, the findings may not be generalisable to EDs with different case mix. Stroke and ST-elevation myocardial infarction requiring reperfusion may be identified as false negative cases in EDs that do not diver these cases to specialist services. Further research is therefore required to reproduce our findings in other settings.

The implications of our findings are that ED staff should avoid over-reliance on NEWS2 in initial assessment. A substantial proportion of patients needing time-critical treatment will have a low NEWS2 score and most patients with NEWS2>4 will not require time-critical treatment. We have identified time-critical interventions that NEWS2 predicts poorly and NEWS2 parameters that may over-predict need for time-critical intervention. Further research is required to confirm these findings in other settings and then explore whether NEWS2 can be modified or augmented to improve prediction.

## Data Availability

Anonymised data are available from the corresponding author upon reasonable request

## Competing interests

All authors have completed the ICMJE uniform disclosure form at www.icmje.org/coi_disclosure.pdf and declare: funding to their employing institutions from the National Institute for Health Research (NIHR) Research for Patient Benefit Programme (project reference NIHR204935); no financial relationships with any organisations that might have an interest in the submitted work in the previous three years; no other relationships or activities that could appear to have influenced the submitted work.

## Contributor and guarantor information

SG conceived the study. SG, LS, GF, RP, EH and LA designed the study. SG acquired the data. SG, GF and AT undertook outcome adjudication. LS analysed the data. All authors interpreted the data. All authors contributed to drafting the manuscript. SG is the guarantor of the paper. The corresponding author attests that all listed authors meet authorship criteria and that no others meeting the criteria have been omitted.

## Acknowledgements

We thank Enid Hirst and Linda Abouzeid (public representatives from Sheffield Emergency Care Forum) for providing the patient and public involvement outlined in the paper, Martin Bayley (Healthcare Computer Scientist, Sheffield Teaching Hospitals NHS Foundation Trust) for providing the hospital data, Erica Wallis (Research Coordinator, Sheffield Teaching Hospitals NHS Foundation Trust) for assistance with research governance and regulatory approvals, and Anna Wilson and Sarah Bird (Research Nurses, Sheffield Teaching Hospitals NHS Foundation Trust) for undertaking the case record screening.

## Data Sharing

Anonymised data are available from the corresponding author upon reasonable request (contact details on first page).

## Funding

The study was funded by the United Kingdom National Institute for Health Research (NIHR) Research for Patient Benefit (RfPB) programme (project reference NIHR204935). The funder played no role in the study design; in the collection, analysis, and interpretation of data; in the writing of the report; and in the decision to submit the article for publication. The views expressed are those of the authors and not necessarily those of the NHS, the NIHR or the Department of Health and Social Care.

## Notes

### Clinical Trial

Research Registry, 10450

### Author Declarations

The United Kingdom Health Research Authority and Health and Care Research Wales reference 23/HRA/4572).

## References

1. Smith E, Higginson I, Cleaver B, Smith M, Morris AM. Initial Assessment of Emergency Department Patients. Royal College of Emergency Medicine, 2017. https://www.rcem.ac.uk/docs/SDDC%20Intial%20Assessment%20(Feb%202017).pdf

2. NHS England 2022. Guidance for emergency departments: initial assessment, published 12^th^ August 2022. https://www.england.nhs.uk/guidance-for-emergency-departments-initial-assessment/ (accessed 02/08/24)

3. Royal College of Physicians. National Early Warning Score (NEWS) 2: Standardising the assessment of acute-illness severity in the NHS. Updated report of a working party. London: RCP, 2017.

4. NHS England Clinical Policy Unit (2019). Resources to support the adoption of the National Early Warning Score. https://www.england.nhs.uk/wp-content/uploads/2019/12/Resources_to_support_the_adoption_of_NEWSFINAL.pdf (accessed 02/09/2024)

5. Arévalo-Buitrago P, Morales-Cané I, Olivares Luque E, Guler I, Rodríguez-Borrego MA, López-Soto PJ. Predictive power of early-warning scores used in hospital emergency departments: a systematic review and meta-analysis. Emergencias. 2021 Oct;33(5):374–381

6. Nannan Panday RS, Minderhoud TC, Alam N, Nanayakkara PWB. Prognostic value of early warning scores in the emergency department (ED) and acute medical unit (AMU): A narrative review. Eur J Intern Med. 2017 Nov;45:20–31.

7. Patel R, Nugawela MD, Edwards HB, Richards A, Le Roux H, Pullyblank A, Whiting P. Can early warning scores identify deteriorating patients in pre-hospital settings? A systematic review. Resuscitiation 2018; 13:101–111.

8. Guan G, Lee CMY, Begg S, Crombie A, Mnatzaganian G (2022) The use of early warning system scores in prehospital and emergency department settings to predict clinical deterioration: A systematic review and meta-analysis. PLoS ONE 17(3): e0265559.

9. Goodacre S. Using clinical risk models to predict outcomes: What are we predicting and why? Emerg Med J 2023;40:728–30.

10. NHS Midlands and Lancashire Strategy Unit 2023. Emergency department acuity measurement and process: quick scoping review. https://www.strategyunitwm.nhs.uk/publications/emergency-department-acuity-measurement-and-process-quick-scoping-review (accessed 02/08/24)

11. Hong R, Sexton R, Sweet B, Carroll G, Tambussi C, Baumann BM. Comparison of START triage categories to emergency department triage levels to determine need for urgent care and to predict hospitalization. Am J Disaster Med. 2015 Winter;10(1):13–21.

12. Hinson JS, Martinez DA, Cabral S, George K, Whalen M, Hansoti B, Levin S. Triage Performance in Emergency Medicine: A Systematic Review. Ann Emerg Med. 2019 Jul;74(1):140–152.

13. Hanley, JA and McNeil, BJ (1982). The meaning and use of the area under a Receiver Operating Characteristic (ROC) curve. Radiology, 148, 29–36.

14. Brown LD, Cai TT, DasGupta A (2001). Interval estimation for a binomial proportion. Statistical Science, 16(2), 101–117.

15. Thomas B, Goodacre S, Lee E, Sutton L, Burnsall M, Loban A et al. Prognostic accuracy of emergency department triage tools for adults with suspected COVID-19: The PRIEST observational cohort study. Emerg Med J 2021; 38: 587–593

16. Hirst E, Irving A, Goodacre S. Patient and public involvement in emergency care research. Emerg Med J 2016;33:665–670.

17. Bion J BG, Boyle A, Carrol E, Christian W, Crossland S, Faust S, et al. Academy of Medical Royal Colleges Statement on the Initial Antimicrobial Treatment of Sepsis. Academy of Medical Royal Colleges; 2022.

