## Supplementary figures and images for "Accuracy of the National Early Warning Score version 2 (NEWS2) in predicting need for time-critical treatment: Retrospective observational cohort study"

### Supplementary figure 1

Supplementary figure 1: Participant flow through the study

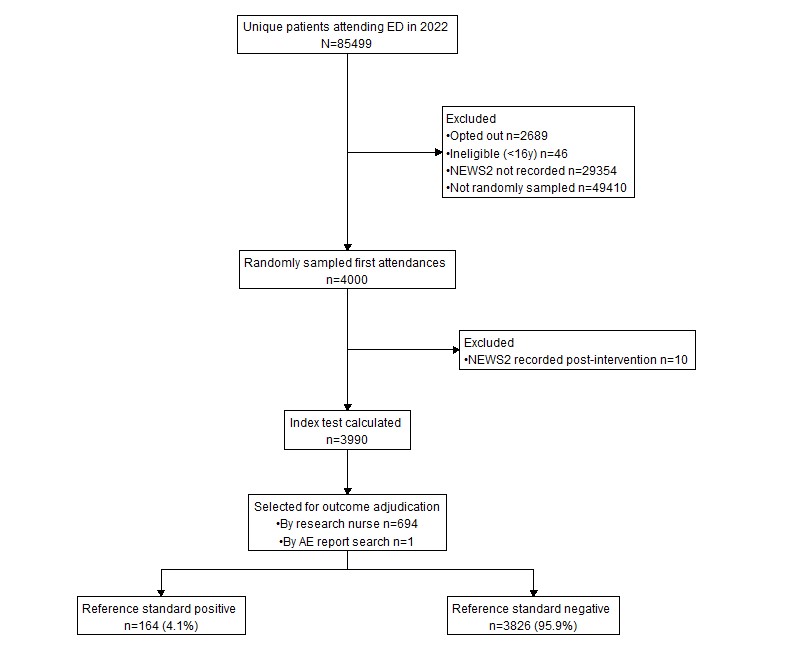
