## Supplementary table 1 for "Accuracy of the National Early Warning Score version 2 (NEWS2) in predicting need for time-critical treatment: Retrospective observational cohort study"

Supplementary table 1: Characteristics of patients with NEWS2≤4 who needed time-critical treatment (N=79)

| **Variable** | **Mean, media, N (%)** |
| --- | --- |
| Age (years) |  |
| Mean (SD) | 58.2 (21.4) |
| Median [Min, Max] | 63.0 [17.0, 92.0] |
| Sex |  |
| Female | 33 (41.8%) |
| Male | 46 (58.2%) |
| Ethnicity |  |
| Asian | 4 (5.3%) |
| Black | 2 (2.7%) |
| Mixed | 1 (1.3%) |
| Other | 2 (2.7%) |
| White | 66 (88.0%) |
| Not stated/NA | 4 |
| Pulse rate (beats/min) |  |
| Mean (SD) | 89.8 (21.5) |
| Median [Min, Max] | 86.0 [58.0, 173] |
| Respiration rate (breaths/min) |  |
| Mean (SD) | 18.5 (2.71) |
| Median [Min, Max] | 18.0 [12.0, 28.0] |
| Oxygen saturation (%) |  |
| Mean (SD) | 97.9 (1.76) |
| Median [Min, Max] | 98.0 [90.0, 100] |
| ACVPU |  |
| Alert | 76 (96.2%) |
| Confused | 3 (3.8%) |
| Voice | 0 (0%) |
| Pain | 0 (0%) |
| Unresponsive | 0 (0%) |
| Systolic BP (mmHg) |  |
| Mean (SD) | 135 (26.7) |
| Median [Min, Max] | 135 [71.0, 197] |
| Diastolic BP (mmHg) |  |
| Mean (SD) | 79.0 (16.0) |
| Median [Min, Max] | 78.0 [47.0, 139] |
| Temperature (C) |  |
| Mean (SD) | 36.6 (0.789) |
| Median [Min, Max] | 36.5 [34.6, 39.1] |
| Missing | 1 (1.3%) |
| Supplemental oxygen |  |
| Air | 67 (84.8%) |
| On O_2_ | 12 (15.2%) |
