## Supplementary table 2 for "Accuracy of the National Early Warning Score version 2 (NEWS2) in predicting need for time-critical treatment: Retrospective observational cohort study"

Supplementary table 2: Characteristics of patients with NEWS2>4 who did not need time-critical treatment (N=245)

| **Variable** | **Mean, media, N (%)** |
| --- | --- |
| Age (years) |  |
| Mean (SD) | 64.6 (22.4) |
| Median [Min, Max] | 71.0 [16.0, 98.0] |
| Sex |  |
| Female | 138 (56.3%) |
| Male | 107 (43.7%) |
| Ethnicity |  |
| Asian | 10 (4.3%) |
| Black | 8 (3.4%) |
| Mixed | 2 (0.9%) |
| Other | 9 (3.9%) |
| White | 204 (87.6%) |
| Not stated/NA | 12 |
| Pulse rate (beats/min) |  |
| Mean (SD) | 103 (26.0) |
| Median [Min, Max] | 101 [18.0, 190] |
| Respiration rate (breaths/min) |  |
| Mean (SD) | 24.1 (6.29) |
| Median [Min, Max] | 24.0 [6.00, 66.0] |
| Oxygen saturation (%) |  |
| Mean (SD) | 95.4 (4.05) |
| Median [Min, Max] | 96.0 [70.0, 100] |
| Missing | 1 (0.4%) |
| ACVPU |  |
| Alert | 186 (75.9%) |
| Confused | 36 (14.7%) |
| Voice | 11 (4.5%) |
| Pain | 7 (2.9%) |
| Unresponsive | 5 (2.0%) |
| Systolic BP (mmHg) |  |
| Mean (SD) | 132 (29.3) |
| Median [Min, Max] | 132 [56.0, 219] |
| Diastolic BP (mmHg) |  |
| Mean (SD) | 77.2 (18.2) |
| Median [Min, Max] | 77.0 [0, 137] |
| Temperature (C) |  |
| Mean (SD) | 37.0 (1.31) |
| Median [Min, Max] | 37.0 [34.0, 40.6] |
| Supplemental oxygen |  |
| Air | 151 (61.6%) |
| On O_2_ | 94 (38.4%) |
