## Supplementary table 3 for "Accuracy of the National Early Warning Score version 2 (NEWS2) in predicting need for time-critical treatment: Retrospective observational cohort study"

Supplementary table 3: NEWS2 parameters scoring three points in patients with NEWS2>4 who did not need time-critical treatment (N=245)

| **NEWS2 parameter** | **Frequency** |
| --- | --- |
| Respiration rate | 95 |
| Supplemental oxygen | 94 |
| ACVPU | 59 |
| Pulse rate | 38 |
| Oxygen saturation | 32 |
| Temperature | 19 |
| Blood pressure | 14 |
