## Supplementary appendix for "Accuracy of the National Early Warning Score version 2 (NEWS2) in predicting need for time-critical treatment: Retrospective observational cohort study"

**Supplementary appendix: Expert consensus development of the outcome measure**

**Background**

The outcome measure, need for time-critical treatment, was defined as either receipt of a time-critical intervention or occurrence of an adverse outcome that could have been prevented by time-critical intervention. To use this outcome measure, we needed to identify a list of potentially time-critical interventions. We therefore assembled a clinical expert group to develop an agreed list of time critical interventions during a three-hour online workshop, informed by the existing evidence base.

**Principles for using the list to determine the outcome**

We intended the list to guide rather than dictate expert adjudication of the outcome, so we used the following principles in developing the list:

1. The list relates to interventions occurring in the emergency department (ED) or shortly after leaving the ED.
2. The list outlines the interventions in general terms rather than specifying the precise treatment and indications.
3. Expert adjudication determines whether the intervention was time-critical and appropriate in each specific case, and therefore whether the intervention meets the outcome definition.
4. The list is not comprehensive but can guide judgements for exceptional cases.
5. Expert adjudication considers ceilings of treatment, such as whether an intervention was or could have been in the patient’s best interests.
6. Expert adjudication considers barriers to intervention, such as whether the intervention received is equivalent to an intervention on the list (e.g. oral or subcutaneous treatment when difficult intravenous access prevents intravenous (IV) treatment).
7. Need for the intervention does not have to be predictable at presentation (e.g. defibrillation for unexpected arrhythmia).

**Background literature**

We identified previous studies that have used need for time-critical treatment as an outcome measure or consensus methods to define need for time-critical treatment. We used these studies to develop the initial list of interventions.

Lerner et al used five informant interviews and a modified Delphi survey to identify fourteen time-specific indicators for major trauma centre need: major surgery, advanced airway, blood products, admission for spinal cord injury, thoracotomy, pericardiocentesis, caesarean delivery, intracranial pressure monitoring, interventional radiology, and in-hospital death.

Challen et al reviewed 25 acute care guidelines to identify a list of 25 potentially life-saving interventions that were then used to classify a primary outcome that included potentially prevented death, in which the patient survived and received a potentially life-saving intervention.

Johansson et al used a Delphi approach involving 18 Swedish triage experts to identify 49 time-critical outcomes (diagnoses, interventions, or events) that could be used when evaluating 5-level priority triage systems. They named this the Lund Outcome Set for Evaluation of Triage (LOSET).

Mercier et al used a modified Delphi study involving 23 multidisciplinary trauma experts to develop a list of 30 trauma care interventions requiring transport to a trauma centre, including endotracheal intubation, blood product administration, angioembolization, intensive care unit admission, and major surgery.

**Initial list of interventions**

We developed an initial list of interventions based on the literature search and clinical expertise in the study team. This included the following interventions:

- Any airway adjunct or procedure to maintain a patent airway.
- Any intervention to support ventilation, other than supplemental oxygen or ventilatory support during sedation.
- Any intervention to support circulation, other than intravenous fluids.
- Unscheduled renal replacement therapy (dialysis or haemofiltration).
- Primary percutaneous coronary intervention or thrombolysis for myocardial infarction.
- Thrombolysis or mechanical thrombectomy for stroke.
- Emergency reperfusion of an ischaemic limb or organ (e.g. testicular torsion).
- Parenteral treatment for hypoglycaemia.
- Insulin infusion for diabetic ketoacidosis or hyperosmolar hyperglycaemic state.
- Hypertonic saline for hyponatraemia.
- Hyperkalaemia or hypokalaemia involving IV treatment and cardiac monitoring.
- IV fluids and steroids for Addisonian crisis.
- IV antibiotics for infection causing new organ dysfunction or shock.
- Administration of blood products.
- Reversal of anticoagulation.
- Any intervention to achieve haemorrhage control, other than manual pressure or a dressing.
- Active rewarming for hypothermia.
- Active cooling for hyperthermia.
- Emergency surgery to avoid death or significant disability.
- Large volume IV fluid replacement (≥2L or ≥1L with central venous monitoring).
- Emergency delivery (vaginal or c-section).
- Cardioversion or rate control for life-threatening arrhythmia.
- Cardiac pacing or IV treatment for bradyarrhythmia.
- Reduction of limb-threatening fracture or dislocation (including threat to skin, nerve or perfusion).
- Adrenaline for anaphylaxis or airway compromise.
- IV antidote for poisoning.
- IV treatment to lower blood pressure.
- IV nitrates for acute heart failure.
- Parenteral sedation for acute behavioural disturbance.
- Pleural decompression for respiratory compromise.
- Pericardial decompression for cardiovascular compromise.

**Expert consensus group review**

We subsequently held a three-hour facilitated round-table expert consensus meeting online on 30^th^ January 2024. Nine emergency medicine clinicians were purposively sampled for their expertise in acute emergency care, to form a consensus clinical panel, ensuring that a range of geographical and hospital settings were represented (district general hospitals and tertiary hospitals, trauma units and major trauma centres). The experts are listed at the end of this appendix.

The literature review findings and initial list of interventions were shared with participants prior to the meeting. They were asked to complete an online survey asking whether each of the initially proposed interventions should be included in the outcome measure. *A priori* thresholds of ≥80% and ≤50% agreement were pre-specified for inclusion and exclusion of each intervention. Respondents also had the opportunity to propose new interventions and suggest caveats or conditions for existing interventions.

The meeting was subsequently chaired by an experienced emergency medicine clinical academic (GF). Brief presentations covered project aims, reference standard principles, expert consensus methodology, and reviewed the pre-meeting survey results. For interventions meeting the threshold for inclusion, additional contextual issues were initially discussed and agreed (e.g., which symptoms represent severe hyponatraemia requiring hypertonic saline). The meeting then proceeded in a structured format using a modified nominal group technique, comprising a round-robin sharing of new interventions, followed by iterative rounds of online anonymous voting on inclusion of interventions (either newly suggested or with no consensus from the pre-meeting survey) and discussion of results (including any caveats). Up to three rounds were planned to achieve consensus. All discussions were facilitated to ensure issues were thoughtfully deliberated, incorporated diverse experience and views, and produced the best possible decision.

**Final list of interventions and supporting documentation**

The expert consensus process resulted in a list of 33 time-critical interventions, with consensus achieved for all interventions after two voting rounds.

Upon implementing the outcome adjudication process, we identified an additional intervention (IV antibiotics for bacterial meningitis or necrotising fasciitis) that was not on the list but clearly should have been included. This was added to the list to give a final list of 34 time-critical interventions, following *post hoc* agreement by the expert panel via email. We subsequently added an open category (labelled intervention #99) that could be used for any unspecified interventions that the outcome adjudicators decided was a time-critical intervention but was not on the list.

Feedback from the consensus group was used to create guidance for outcome adjudicators around assessment of prehospital interventions, withdrawal of treatment or futility, inappropriate use of interventions, and specification of indications for the interventions. This guidance was further developed during the study by drawing on the experience of the outcome adjudicators using the list.

The final list and associated guidance are outlined below.

**Final list of interventions**

1. Unplanned airway intervention to provide a patent airway, other than during sedation for a procedure.
2. Any intervention to support ventilation (except supplemental oxygen), other during sedation for a procedure.
3. Any intervention to support circulation (including CPR), other than intravenous fluids.
4. Unscheduled renal replacement therapy (dialysis or haemofiltration).
5. Primary percutaneous coronary intervention or thrombolysis for myocardial infacrtion.
6. Thrombolysis or mechanical thrombectomy for stroke.
7. Thrombolysis for massive pulmonary embolism.
8. Emergency reperfusion of an ischaemic limb or organ (e.g. testicular torsion).
9. Parenteral treatment for hypoglycaemia.
10. Insulin infusion as part of treatment protocol for diabetic ketoacidosis or hyperosmolar hyperglycaemic state.
11. Hypertonic saline for hyponatraemia causing a specific neurological disturbance, such as seizures or reduced conscious level.
12. Hyperkalaemia or hypokalaemia involving IV treatment and cardiac monitoring.
13. IV fluids and steroids for Addisonian crisis.
14. IV antibiotics for infection causing new organ dysfunction or shock.
15. Administration of blood products for acute blood loss, to allow emergency procedures, in haematological emergencies, or severe anaemia in context of proven acute coronary syndrome.
16. Reversal of anticoagulation to prevent loss of life or significant disability or to facilitate a procedure or surgery.
17. Any intervention to achieve haemorrhage control, other than manual pressure or a dressing.
18. Active rewarming for hypothermia.
19. Active cooling for hyperthermia.
20. Emergency surgery to avoid death or significant disability, including surgical source control for infection.
21. Large volume IV fluid replacement (>2L within 2 hours or >1L with central venous monitoring).
22. Emergency delivery requiring healthcare professional intervention for mother or baby.
23. Cardioversion or rate control for life-threatening arrhythmia.
24. Cardiac pacing or IV treatment for bradyarrhythmia.
25. Reduction of limb-threatening fracture or dislocation (including threat to skin, nerve or perfusion).
26. IV antibiotics for genuine open fractures.
27. Adrenaline for anaphylaxis or airway compromise.
28. Antidote for life or disability threatening poisoning.
29. IV treatment to lower life or disability threatening blood pressure.
30. IV nitrates for acute heart failure.
31. Parenteral sedation for acute behavioural disturbance.
32. Pleural decompression for respiratory compromise.
33. Pericardial decompression for cardiovascular compromise.
34. IV antibiotics for meningitis or necrotising fasciitis.

99. Any other, not previously specified

**Guidance for outcome adjudicators**

Prehospital interventions:

Only include ED interventions. If a prehospital intervention has been completed on arrival at the ED (e.g. fracture reduced, arrhythmia cardioverted), then identification of time-critical intervention starts from arrival at the ED. If a prehospital intervention is ongoing on arrival at the ED (e.g. airway adjunct or ventilatory support), then assess whether it still represents an urgent treatment (i.e. would it need to be provided urgently if it had not already been done). If the prehospital intervention no longer appears to be needed (e.g. airway adjunct post-seizure or post-respiratory arrest that could be removed), then no time-critical ED intervention has occurred.

Withdrawal of treatment or futility:

Include interventions that were withdrawn or deemed to be futile after a trial of treatment. Exclude interventions that were withdrawn or deemed to be futile on the basis of additional information that was not available at the time the intervention was given (such as patient preference or clinician expertise).

Inappropriate interventions:

Do not include interventions that were clearly not indicated but use a high threshold for applying this judgement. This should be based on guidance specifying the indications for the intervention, rather than personal preference.

#2 Any intervention to support ventilation

Include patients using their own non-invasive ventilation if it was used to treat new or worsening type 2 respiratory failure rather than just continuation of home treatment.

#5 Primary percutaneous coronary intervention or thrombolysis for myocardial infarction

Do not include non-ST elevation coronary syndromes unless they require emergency percutaneous coronary intervention for ongoing pain and dynamic ST changes.

#12 Hyperkalaemia or hypokalaemia involving IV treatment and cardiac monitoring

If use of cardiac monitoring is not recorded or appears inappropriate, include if potassium is given at ≥10mmol/h for hypokalaemia (<3mmol/L) or if calcium chloride/gluconate is given for hyperkalaemia (>6mmol/L).

#13 Intravenous fluids and steroids for Addisonian crisis

Do not include if IV fluids and steroids were given for people thought to be steroid dependent but without features of Addisonian crisis.

#14 Intravenous antibiotics for infection causing new organ dysfunction or shock

Definition of infection causing new organ dysfunction or shock:

1. Microbiological or radiological evidence of an infection that can cause organ dysfunction and could be effectively treated by the relevant antimicrobial agent.
2. Evidence of shock or new respiratory, renal, hepatic, or bone marrow failure recorded within 24h of presentation.

Shock is defined as persisting hypotension (mean arterial pressure <65mmHg or use of vasopressors to maintain mean arterial pressure above 65mmHg) and serum lactate >2 mmol/L despite adequate volume resuscitation.

New organ dysfunction must be primarily due to the infection, so it does not include:

1. Abnormal liver function tests associated with cholecystitis (i.e. cases where there is no clear evidence of infection, such as bacteraemia or a collection or gas in the biliary tree on imaging). Abnormal liver function tests due to ascending cholangitis is included.
2. Respiratory failure primarily due to exacerbation of chronic lung disease or non-infective lung pathology. Respiratory failure primarily due to pneumonia is included.
3. Pre-renal acute kidney injury primarily due to fluid deficiency or post-renal acute kidney injury primarily due to obstruction. This judgement may be assisted by assessing how the acute kidney injury responds to improving fluid balance. A rapidly corrected acute kidney injury is unlikely to be primarily due to infection.

#17 Any intervention to achieve haemorrhage control

Include nasal packing for significant uncontrolled epistaxis if pressure was clearly unsuccessful or inappropriate to control the bleeding. Include wound suturing if pressure or dressing was clearly unsuccessful or inappropriate to control the bleeding.

#20 Emergency surgery

The inclusion criteria for the National Emergency Laparotomy Audit can determine whether a laparotomy was emergency surgery (<https://www.nela.org.uk/Criteria>) but this only applies to general surgery laparotomy (not gynaecological). Include pleural and pericardial procedures (aspiration, drain) if required to treat respiratory failure or circulatory shock.

#21 Large volume IV fluid replacement

Determine from the fluids prescribed rather than fluids given. Assume that fluids prescribed as ‘stat’ are effectively given in zero time. There must be a reasonable indication for large volume IV fluids but, as above, use a high threshold if excluding this intervention as inappropriate.

#23 Cardioversion or rate control for life-threatening arrhythmia

As per Resuscitation Council Advanced Life Support guidance, this should be judged on evidence of shock, syncope, ischaemic chest pain, or severe heart failure.

#25 Reduction of limb-threatening fracture or dislocation

Limb-threatening can be determined if the notes or discharge summary record a threat to limb (tight skin, neurovascular compromise) or state that urgent manipulation was required, if a radiology report notes potentially limb-threatening features, or if the limb was manipulated before x-ray.

#29 IV treatment to lower life or disability threatening high blood pressure

This includes lowering blood pressure for a specific cause, such as intracerebral haemorrhage or acute aortic syndrome.

**References**

Lerner EB, Willenbring BD, Pirrallo RG, Brasel KJ, Cady CE, Colella MR, Cooper A, Cushman JT, Gourlay DM, Jurkovich GJ, Newgard CD, Salomone JP, Sasser SM, Shah MN, Swor RA, Wang SC. A consensus-based criterion standard for trauma center need. J Trauma Acute Care Surg. 2014 Apr;76(4):1157-63.

<https://pubmed.ncbi.nlm.nih.gov/24662885/>

prevented death, where the patient survived and received a potentially life-saving intervention.

Challen K, Bradburn M, Goodacre SW. Development and validation of a score to identify in the Emergency Department patients who may benefit from a time-critical intervention: a cohort study. Scand J Trauma Resusc Emerg Med. 2015 Sep 17;23:67.

<https://pubmed.ncbi.nlm.nih.gov/26383093/>

Johansson A, Ekwall A, Forberg JL, Ekelund U. Development of outcomes for evaluating emergency care triage: a Delphi approach. Scand J Trauma Resusc Emerg Med. 2023 Feb 25;31(1):10.

<https://www.ncbi.nlm.nih.gov/pmc/articles/PMC9958312/>

Mercier É, Nadeau A, Le Sage N, Moore L, Malo C, Blanchard PG, Fleet R, Émond M. A Canadian consensus-based list of urgent and specialized in-hospital trauma care interventions to assess the accuracy of prehospital trauma triage protocols: a modified Delphi study. Can J Surg. 2023 Mar 31;66(2):E181-E188.

<https://www.ncbi.nlm.nih.gov/pmc/articles/PMC10069413/>

**Experts involved in the consensus meeting**

Fiona Lecky

Tim Coats

Andreas Crede

Ed Carlton

Andrew Tabner

Graham Johnson

Alasdair Gray

Matthew Reed

Kirsty Challen
